# How should doctors manage workplace interruptions? A qualitative study in UK hospital outpatient departments

**DOI:** 10.64898/2026.10.14.26365522

**Authors:** Shaun Boustani, Chase Staras, Antonia Rich, Ann Griffin

**Author notes:** **Correspondence to** Dr Shaun Boustani, UCL Research Department of Medical Education, UCL Medical School, University College London, London, UK.

## Abstract

**Background:** Interruptions are common in hospital outpatient departments (OPDs) and are linked to error. Many are clinically necessary, so the aim is to regulate them. Most interventions target the individual clinician. Little is known about how doctors manage interruptions in practice or what makes their strategies work.

**Methods:** Qualitative study using focus groups and interviews with 42 doctors working in UK National Health Service OPDs (19 sessions, September to November 2025). Participants were purposively sampled by clinical experience and self-reported patient complexity. Data were analysed using Framework Analysis, informed by Cognitive Load Theory and Social Identity Theory.

**Results:** We developed four themes. (1) *Pre-emptive protection of attention*. Doctors withdrew, signalled unavailability and set thresholds for interrupting. These depended on colleagues learning and sharing the norm, which staff rotation eroded. (2) *Sharing the burden through team coordination*. Designated roles and time-limited rotas moved interruptions to colleagues with capacity set aside for them and made interrupting legitimate. (3) *Real-time cognitive repair*. Doctors asked interrupters to wait while they secured the task, and rebuilt the consultation on return, sometimes with the patient’s help. Asking a colleague to wait depended on seniority or team membership; some trainees described difficulty doing this with senior colleagues. (4) *Organisational recognition as the precondition for sustainable regulation*. Where supervision was built into job plans, the interruptions it generated were expected and doctors held capacity in reserve for them. Advice, vetting and telephone queries were rarely counted as work.

**Conclusions:** Each strategy served a cognitive purpose, and social and organisational conditions decided whether it was perceived to work. Interventions could make norms shared and roles explicit, count interruption work in job plans and keep routes open for junior staff to raise concerns. Evaluations should ask who is able to use them.

**WHAT IS ALREADY KNOWN ON THIS TOPIC:** Interruptions are common in clinical work and can disrupt task completion, while most interventions focus on individual clinicians and have shown mixed effects.

**WHAT THIS STUDY ADDS:** Doctors described strategies to protect attention, redistribute interruptions and repair disrupted tasks, but their use depended on shared norms, legitimate roles, social standing and organisational capacity. Some trainees described difficulty asking senior colleagues to wait, although trainee representation in this sample was limited.

**HOW THIS STUDY MIGHT AFFECT RESEARCH, PRACTICE OR POLICY:** Departments could agree urgency thresholds alongside clear routes for non-urgent requests, use time-limited roles for queries and recognise advice and supervision in workload planning. Evaluations should assess who can use these arrangements and whether staff can still escalate concerns.

## Introduction

Hospital outpatient departments (OPDs) provide most specialist assessment and follow-up in the NHS. In 2024–25 the NHS in England delivered 146.1 million outpatient appointments,^1^ and OPDs carry key responsibility for national targets on waiting times and cancer care.^2^ Work interruptions are unexpected suspensions of attention or activity on an ongoing task.^3^ Across clinical work they are linked to higher workload, errors and tasks left unfinished.^4–6^ Laboratory studies place much of their cost in resuming the interrupted task.^7–9^

Many interruptions are also necessary. They carry advice, supervision, escalation of concern and information that keeps patients safe.^10^ A review of nursing research found little evidence that reducing interruptions improves patient outcomes.^11^ The practical question is how to regulate interruptions while keeping the useful ones.

Most interventions target the individual clinician. Do-not-interrupt signs, tabards and protected time have shown modest and inconsistent effects, mainly during medication administration.^12,13^ Structured communication tools have a growing evidence base at team level,^14^ but team coordination arrangements and organisational measures have received little formal evaluation. Human factors research has described the strategies individuals use, such as suspending the primary task, multitasking, mediating resumption or blocking the interruption.^15^ It has also described how interruptions can be coordinated, whether immediately, by negotiation, through a mediator or by scheduling.^16^ This work tells us what individuals do. It says little about the team and organisational conditions that make a strategy usable, although interruptions are increasingly understood as sociotechnical events involving several people and systems.^17^

These conditions may matter especially in OPDs. Trainees rotate through clinics often,^18^ and our companion paper, drawn from the same dataset, found that agency staff and cross-specialty cover may add to this churn. The colleagues a doctor interrupts, and is interrupted by, may therefore be unfamiliar. The same paper found that transient teams may weaken the norms that regulate interrupting, and that approachability and hierarchy was felt to concentrate the burden on particular doctors.^19^ The present paper turns from the burden to how doctors manage it.

We drew on two theories. Cognitive Load Theory (CLT) holds that working memory is limited, so an interruption competes for capacity with the task it suspends, and the cost grows with the complexity of that task.^20^ Social Identity Theory (SIT) holds that people sort themselves and others into groups by role, grade and team, and that these groups shape trust, communication and norms.^21–23^ People tend to favour their own group, and shared group norms define what counts as acceptable conduct.^22^ In hospital medicine, boundaries of specialty and grade shape how doctors respond to colleagues from other groups.^24^ SIT therefore suggests that who may interrupt whom, and who may ask for a pause, will follow these boundaries. We used CLT to ask what each strategy does for the clinician’s limited capacity, and SIT to ask which social conditions allow a strategy to be used. We asked:

1. what strategies doctors use to manage interruptions in NHS OPDs
2. how cognitive demands and social conditions, such as familiarity, hierarchy and team stability, shape doctors’ use of interruption-management strategies and the conditions under which these strategies are perceived to be usable.

## Methods

### Design

We conducted a qualitative study using focus groups and individual interviews. Reporting follows the Consolidated Criteria for Reporting Qualitative Research (COREQ).^25^ This paper and its companion report different research questions from one dataset. The companion paper examines how the burden of interruptions is distributed among doctors, whereas this paper examines the strategies doctors use to manage interruptions and the cognitive, social and organisational conditions that enable or constrain those strategies.^19^

### Participants and sampling

Participants were doctors working in NHS hospital OPDs in the UK. Respondents to an online screener survey, which formed part of a national survey of UK secondary care doctors,^26^ were stratified into six groups by clinical experience (trainee, 0–19 years post-qualification, or 20+ years) and self-reported patient complexity (high or low), as detailed in online supplemental file 1. CLT and prior research identify both factors as shaping the cognitive impact of interruptions.^27,28^ Of 207 eligible respondents, 42 doctors took part across 19 sessions (13 focus groups and 6 interviews), representing a broad range of specialties (table 1). Recruitment continued until the sample was judged to hold sufficient information power for the study aims.^29^ No participant withdrew. All names are pseudonyms.

**Table 1.** Participant characteristics (n=42)

| Characteristic | n (%) | Data collection sessions |
| --- | --- | --- |
| **Sampling group (experience; patient complexity)** |  |  |
| G1 Trainee; high | 3 (7.1) | 3 interviews |
| G2 Trainee; low | 2 (4.8) | 2 interviews |
| G3 0–19 years; high | 10 (23.8) | 3 focus groups |
| G4 0–19 years; low | 5 (11.9) | 1 interview; 2 focus groups |
| G5 20+ years; high | 13 (31.0) | 1 interview; 3 focus groups |
| G6 20+ years; low | 9 (21.4) | 1 interview; 3 focus groups |
| **Specialty** |  |  |
| Medicine (physicians) | 11 (26.2) |  |
| Emergency medicine | 8 (19.0) |  |
| Surgery | 7 (16.7) |  |
| Obstetrics and gynaecology | 4 (9.5) |  |
| Paediatrics | 3 (7.1) |  |
| Radiology | 3 (7.1) |  |
| Psychiatry | 2 (4.8) |  |
| Anaesthetics | 1 (2.4) |  |
| Ophthalmology | 1 (2.4) |  |
| Other | 2 (4.8) |  |
| **Years of clinical experience**, mean (SD) | 20.4 (9.7) |  |
| **Self-rated interruption frequency**, mean (SD)* | 4.0 (1.0) |  |

\*Rated on a 5-point scale from 1 (low) to 5 (high).

### Data collection

Sessions were held online via Microsoft Teams between September and November 2025 and lasted about 55 minutes. Focus groups were grouped by experience level to limit hierarchical dynamics. They were led by SB and supported by CS, using a semi-structured topic guide covering interruption types, cognitive and social impacts, and management (online supplemental file 2). When only one participant signed up for a session, it was run as an interview by a single researcher (SB or CS) to reduce any perceived power imbalance. Sessions were audio-recorded, transcribed automatically and checked against the recordings. Analytical memos were written during and immediately after each session. We asked about outpatient work, but participants also drew on emergency, theatre, ward and reporting settings when describing strategies. We kept these accounts because the same doctors work across settings, and the text notes where a strategy came from another setting.

### Analysis

Data were analysed using Framework Analysis^30^ in NVivo 14, moving through familiarisation, coding, development and application of an analytical framework, charting and interpretation. Four researchers with backgrounds in cognitive psychology, social psychology, clinical medicine and health psychology developed a hierarchical codebook. It organised codes into five domains (nature of interruptions, cognitive factors, social dynamics, the intersection of cognitive and social factors, and management practices), and open coding was kept for new material. This paper reports the management practices domain, read alongside related cognitive and social codes. The companion paper reports the other four domains.^19^ One researcher (CS) coded all transcripts and a second (AR) independently coded 10%.

Agreement was high (Cohen’s κ=0.97), which we used as a check that the codebook was applied consistently. Themes were developed from the coded data and then interpreted through CLT and SIT. Draft findings, including the management themes, were shared with all participants for validation (online supplemental file 1).^31^ All 11 written responses agreed that the findings reflected their experience, and one added that interruptions built up across a clinical session.

### Reflexivity

The two facilitators, SB and CS, both men, had backgrounds in cognitive and social psychology. These drew their attention to task switching and expertise, and to interpersonal and organisational dynamics, respectively. The other two researchers, both women, brought clinical and health psychology perspectives. AG is a medical doctor and AR is a health psychologist. Team discussion kept cognitive and social explanations in view together, and we looked for accounts that complicated emerging interpretations. Researchers had no prior relationship with participants, who were told only that the study concerned interruptions and their management.

### Patient and public involvement

Patients and the public were not involved in the design, conduct, reporting or dissemination of this research.

### Ethics

Ethical approval was granted by the UCL Research Ethics Committee on 20 May 2025 (ID 1132). Participants gave written consent via the screener survey and verbal consent to recording at the start of each session.

## Results

We developed four themes (table 2). Read in sequence, they describe how doctors tried to prevent, redirect and repair interruptions, and the organisational conditions that enabled these strategies. The first three themes describe strategies. The fourth describes a condition that participants saw as necessary for those strategies to last. Quotations labelled Q1 to Q13 appear in table 3, and full versions of quotations given in part are in online supplemental file 3.

**Table 2.**
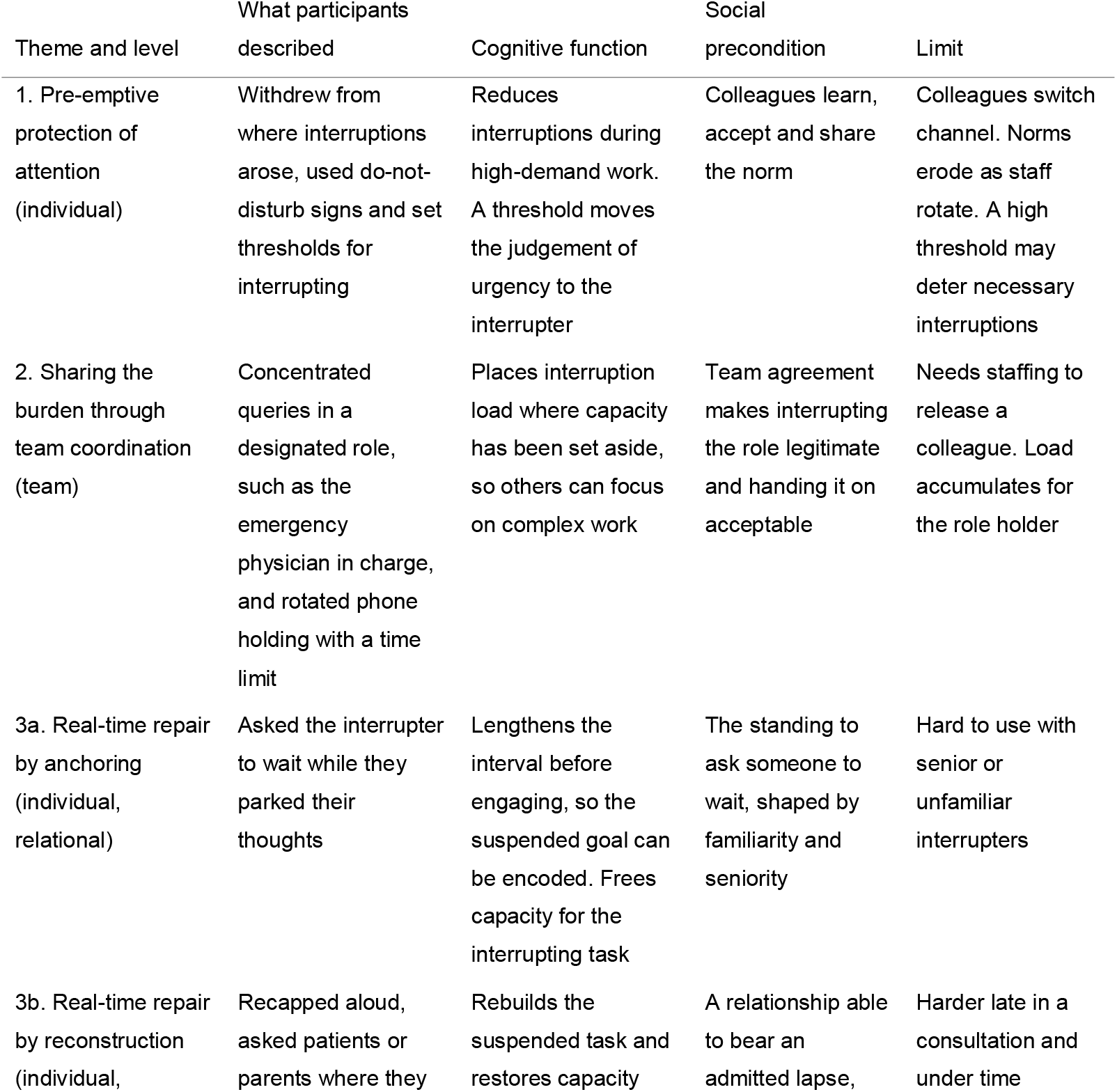
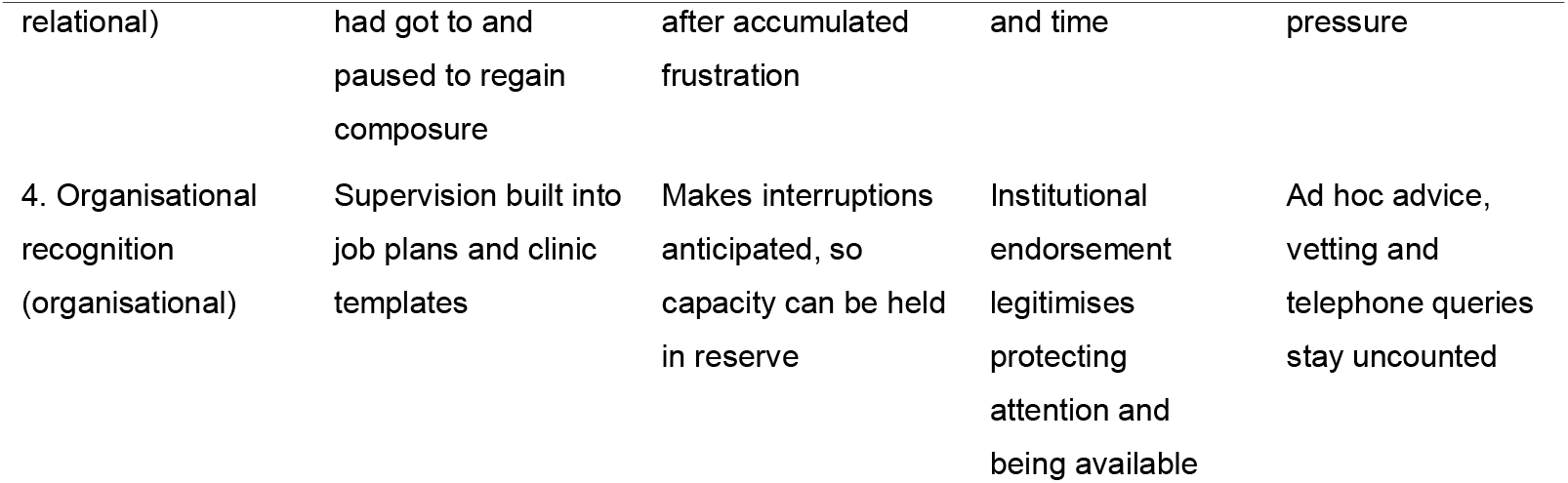
Strategies and conditions for managing interruptions, by theme.

**Table 3.**
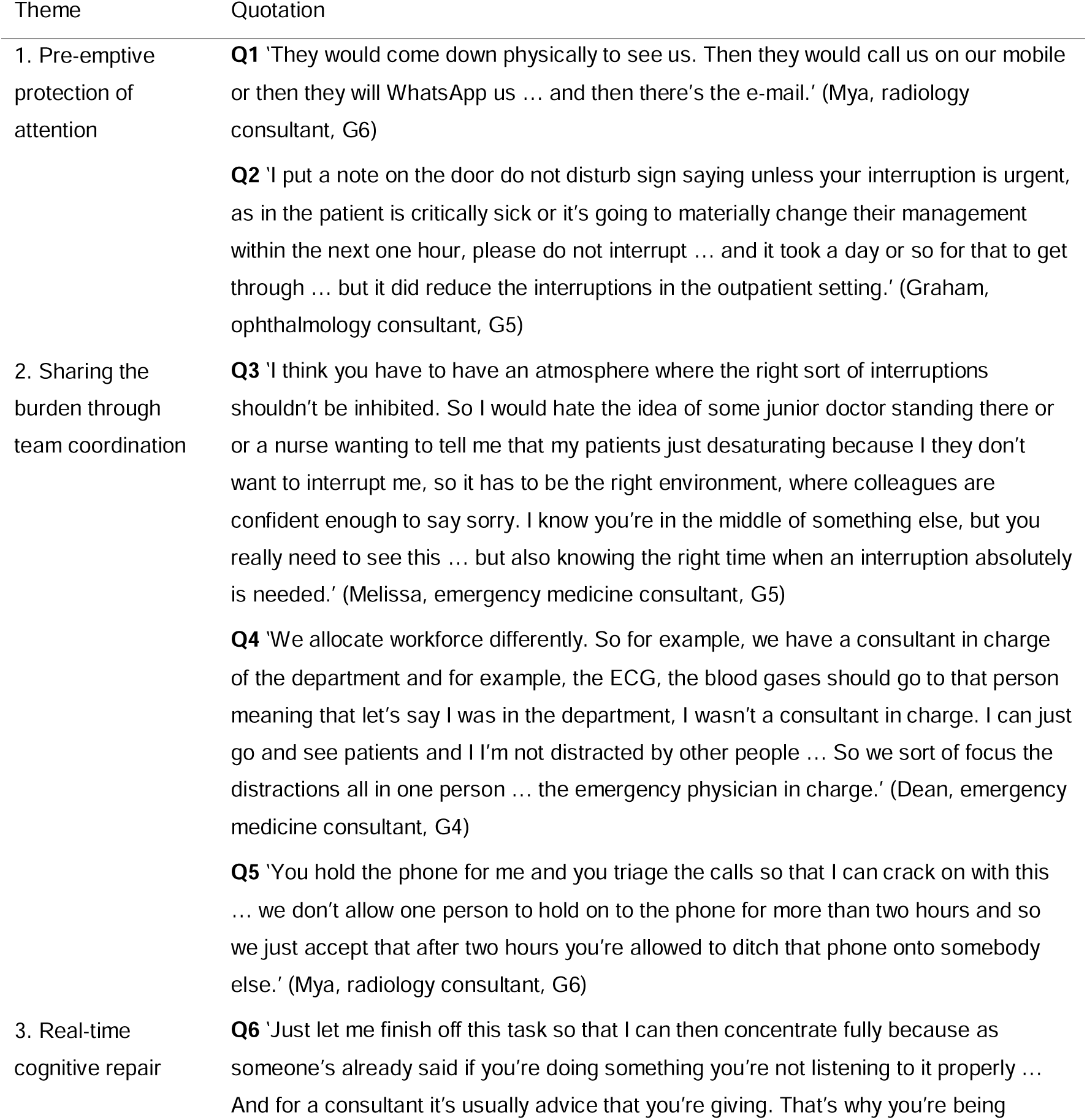
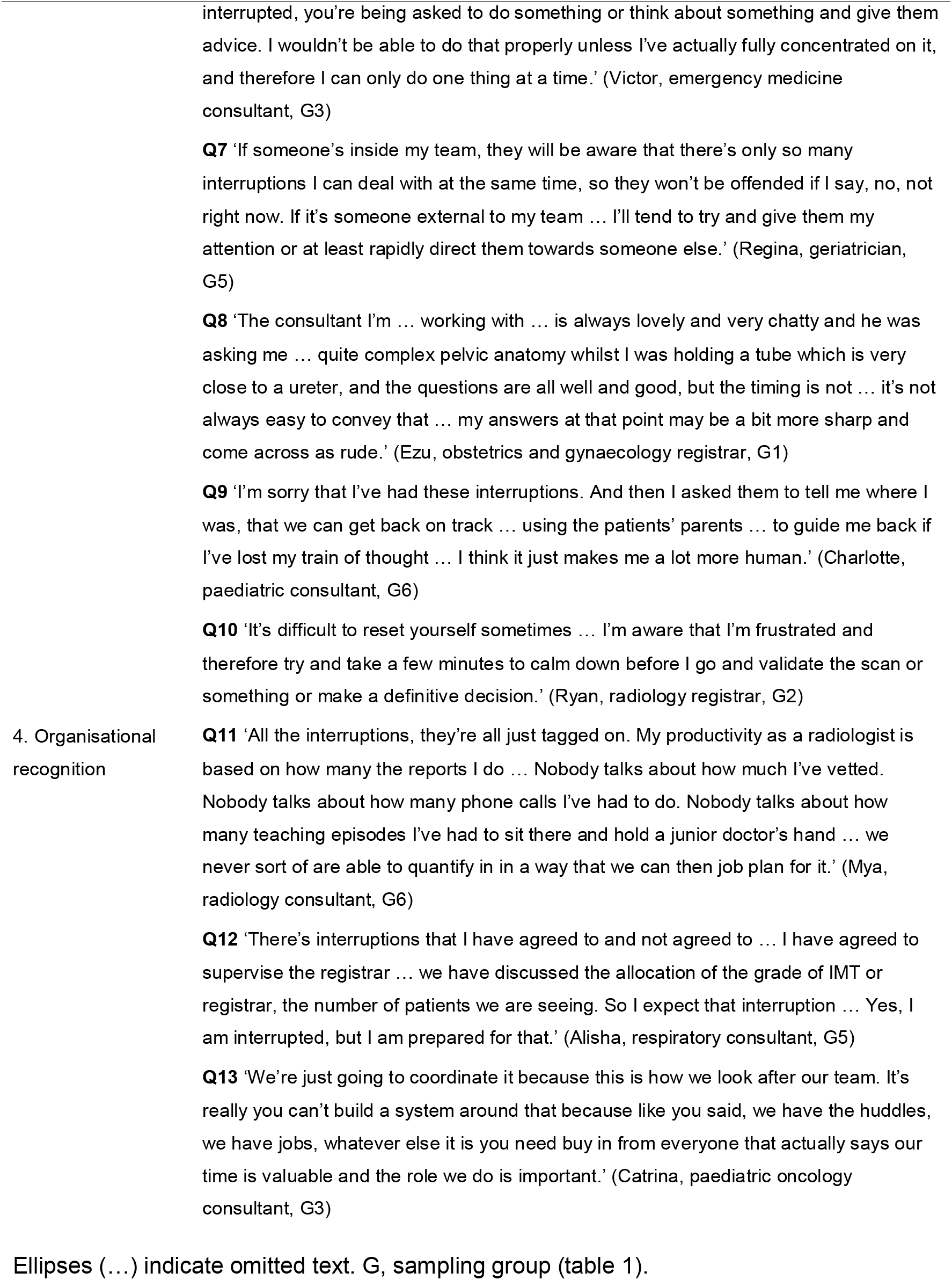
Illustrative quotations.

### Theme 1. Pre-emptive protection of attention

Doctors tried to avoid being interrupted during demanding work. These were individual workarounds without organisational endorsement. Participants described a progression from withdrawing, to signalling unavailability, to specifying when interruption was warranted.

The most common strategy was to become harder to find, to ‘secrete myself in a corner somewhere out of the way’ (Dean, emergency medicine consultant, G4). Withdrawal assumed that interruptions arrived in person. Participants described colleagues switching channel instead, pursuing one request by visit, mobile phone, WhatsApp and email in turn (Q1). Each new channel produced a fresh interruption for the same query.

Some doctors signalled that they should not be disturbed. A generic notice, such as a hotel ‘do not disturb’ sign, ‘doesn’t seem to deter people’ (Graham, ophthalmology consultant, G5). It told colleagues the doctor was busy, but left each of them to decide whether their own request justified an exception. The same participant described a revised sign that set a threshold, a critically ill patient or a change in management within the hour, and reported that it reduced interruptions after a day or so (Q2). Another participant described a colleague’s sign that asked interrupters to confirm they could state the problem clearly and had already consulted their supervisor. In cognitive terms, both signs asked the interrupter to judge urgency before knocking, which spared the interrupted doctor that judgement mid-task.

These strategies depended on colleagues. A threshold took time to take effect and held only once colleagues had learned and accepted it. With trainees rotating every few months, that shared understanding could not be assumed to last. The limit lay in what one doctor could not control alone, namely how colleagues chose to make contact and whether they shared the doctor’s expectations.

### Theme 2. Sharing the burden through team coordination

Team strategies brought colleagues into the solution. Participants set one condition. Any arrangement had to protect attention without discouraging the interruptions that matter, such as a junior doctor or nurse reporting that a patient is deteriorating (Q3).

Some teams concentrated interruptions in a designated role. In one emergency department, results and queries went to the emergency physician in charge (EPIC), so other consultants could see patients without being ‘distracted by other people’ (Q4). Participants described two effects. The role removed interruptions from colleagues doing complex work. It also made approaching the EPIC a proper use of the role, which participants contrasted with imposing on a busy senior. Because the arrangement was agreed at team level, it did not depend on each colleague learning each doctor’s preferences.

For the role holder, interruptions became the task, and Dean described a working memory saturated by the end of the shift. Teams therefore rotated the role. In one radiology department, no one held the phone for over two hours (Q5). Handing it on was something ‘we just accept’, so passing the phone carried none of the social cost of refusing an interruption. The permission came from a norm the team held in common, so no one had to claim it alone.

Team strategies reallocated interruptions. They placed load where capacity had been set aside and made it legitimate both to interrupt and to hand the load on. They depended on staffing that could release one colleague for the role,^4^ and some interruptions still reached the individual doctor however well a team was coordinated.

### Theme 3. Real-time cognitive repair

When interruptions could not be prevented or redirected, doctors managed their cost as it occurred. They anchored the task before engaging and reconstructed it on return.

#### Anchoring before engaging

Doctors’ first priority was to secure the current task. One consultant asked interrupters to ‘hang on while I finish typing the sentence’, which ‘makes all the difference, because it means I can at least park my thoughts before they start speaking to me’ (Jeanne, anaesthetics consultant, G3). Parking her thoughts left the task in a state that could be resumed and freed capacity for the interruption. For consultants, the interruption was often itself a request for advice that needed full attention (Q6).

Anchoring had two preconditions. The first was cognitive. It depended on having ‘enough presence of mind’, which draws on the same capacity it protects. The second was social, because anchoring meant asking someone to wait. The standing to make that request came from two sources. One was shared team membership. With her own team, Regina, a consultant, could say ‘not right now’ without causing offence. With someone from outside the team she would give her attention or redirect them (Q7). The other was seniority. Ezu, an experienced registrar, described a consultant asking complex anatomy questions in theatre while she held a tube close to a ureter. The timing was poor, but she found this hard to convey, and her answers risked coming across as ‘rude’ (Q8). She divided her attention between the procedure and the question.

#### Reconstructing the clinical narrative

On return, doctors rebuilt the interrupted task, often aloud with the patient present. Katrina summarised the history back to the patient ‘just to give my brain time to like get back into what we’re doing’ (Katrina, emergency medicine senior registrar, G2). Others asked patients or parents to say where the consultation had got to (Q9). This made the patient a store for what the doctor had lost. It also meant admitting a lapse, which Charlotte felt made her ‘a lot more human’. We read this as depending on a relationship able to bear the admission.

Reconstruction was harder late in a consultation, when ‘there isn’t enough time to catch up’ (Ben, cognitive disorders specialist, G5). It also had an emotional side. One registrar described taking a few minutes to calm accumulated frustration before validating a scan or making a definitive decision (Q10).

Across both subthemes, repair was cognitive in function and social in its preconditions. It also needed time and spare capacity, which depended on staffing and workload set at organisational level.

### Theme 4. Organisational recognition as the precondition for sustainable regulation

Participants saw sustainable management as depending on time, staffing and recognition that interruption work happened at all. Advice, teaching and the labour of managing interruptions were absent from how workload was measured. One radiologist described productivity judged by reports completed, with vetting, telephone calls and teaching left uncounted, so that ‘we never sort of are able to quantify in in a way that we can then job plan for it’ (Q11).

Participants with and without recognised supervision described the same kind of interruption differently. Macy described clinics that left ‘enough time to see your own patient, but they expect you to do training on top of that’ (Macy, consultant, G3). Each trainee query competed with a full list. Alisha’s supervision had been agreed in advance, including the trainee’s grade and the number of patients booked (Q12). For her, supervisory interruptions were expected work, and she held capacity in reserve for them. This contrast suggests that organisational recognition may change the cognitive cost of an interruption, as well as justifying resources.

Participants also described local etiquette about when to interrupt, and saw it as fragile without wider endorsement (Q13). Organisational recognition did two things that individuals and teams could not do alone. It reserved capacity for interruption work, and it legitimised both protecting attention and being available.

### Differences across sampling groups

The clearest difference was by seniority. Consultants described anchoring as something they could request, while trainees described needing a pause they could not easily ask a senior for. Consultants also lost this standing with colleagues from outside their team (Q7).

Accounts of organisational recognition came mainly from consultants with supervisory roles. We saw no clear differences in strategy use by self-reported patient complexity. Few trainees took part, so the seniority pattern rests on a relatively smaller number of accounts.

## Discussion

### Principal findings

Doctors described strategies that protected attention in advance, redistributed interruptions across teams and repaired their cognitive cost in real time, all within organisational conditions largely beyond their control. Each strategy served a cognitive purpose, protecting limited working memory. Whether it worked depended on social conditions. These were whether colleagues shared a norm, whether a role made interrupting legitimate, whether a doctor had the standing to ask someone to wait and whether the organisation recognised the work at all.

This paper contributes by offering a new catalogue of interruption-management strategies and an explanation of why familiar strategies are not equally available in practice. Individual strategies depended on shared norms, and sustaining them required organisational recognition of interruption work. This helps explain why interventions focused on individual behaviour alone may be insufficient, even when the underlying strategy is cognitively sensible.

### How this extends accounts of interruption handling

The strategies themselves are familiar. In Colligan and Bass’s account of nurses’ strategies, anchoring corresponds to mediating resumption, withdrawal and signs to blocking, and divided attention to multitasking.^15^ Emergency physicians describe similar cognitive strategies.^32^ In McFarlane and Latorella’s terms,^16^ asking a colleague to wait is negotiated coordination, the EPIC and the phone holder are mediated coordination, and supervision built into a job plan is scheduled coordination. Our findings add the conditions under which each form is available. Negotiation needs standing, mediation needs staffing and a shared norm, and scheduling needs organisational planning.

### Why individual strategies reach a ceiling

Do-not-interrupt measures have had mixed effects. In one hospital programme, patients interrupted nurses wearing tabards less often, but staff did not change their behaviour.^33^ Results like this are often read as a problem of compliance. Our findings suggest that the design of the signal also matters. Coiera argues that interruptions persist because they are locally rational for the interrupter, who gains immediate resolution while the cost falls on the person interrupted.^34^ A generic sign leaves this asymmetry intact. It hands the decision about exceptions to the person with most reason to make one. The signs our participants found useful specified a threshold, which moved the triage of urgency from the interrupted doctor to the interrupter. Even so, they worked only once colleagues shared the norm, and in rotating teams the norm must be rebuilt with each cohort. SIT suggests why. Norms draw their force from group membership, which one doctor cannot create alone.^22^ Our national survey points the same way, as individual strategy use was not consistently associated with lower mental workload.^26^

Specified thresholds also carry a risk. They move the judgement of urgency to the interrupter, who is often a junior or unfamiliar colleague. Our companion paper found that concerns raised by such colleagues already risk being discounted.^19^ A threshold set too high could add a further barrier to the interruptions participants most wanted to keep (Q3). Pairing a threshold with a named alternative route, such as the EPIC or the phone holder, gives non-urgent requests somewhere to go.

### Team coordination reallocates interruptions

Collaborative extensions of CLT propose that groups can distribute cognitive processing across members when coordination costs are low.^35^ The EPIC role is one example, in which one clinician absorbs the interruption load so that others can reserve capacity for complex work. For the role holder, the cost shifts from resumption to accumulated saturation, which is why rotation was part of these arrangements. A designated role also changes the social meaning of interrupting. Hierarchy deters junior staff from speaking up,^36,37^ and approaching a role holder is a legitimate use of the role. Because the norm is held by the team, it does not depend on colleagues knowing one another, which rotating OPD teams often do not. However, familiarity still matters. In trauma teams, shared experience strengthens the team’s knowledge of who knows what, and this has been linked to better patient outcomes.^38^ Designated roles may offer a partial substitute where that familiarity is missing. The roles our participants described came from emergency and radiology departments, which already have coordinating posts. Whether outpatient clinics could use an equivalent, such as a named clinician for clinic queries, needs testing. Evaluations of such roles should measure whether staff remain able to escalate concerns, as well as how often interruptions occur.

### Real-time repair is socially licensed

Doctors’ repair strategies match experimental work on task resumption. A suspended task is resumed more quickly when its goal is encoded during the interval before engaging with the interruption.^8,39^ Asking an interrupter to ‘hang on’ lengthens that interval. Participants’ sense that late interruptions were harder to recover from fits evidence that interruptions during high-workload moments are especially costly.^40^

Experimental paradigms hold the social situation constant. In practice, lengthening the interval means asking another person to wait, and whether that is possible depends on who is asking whom. Our companion paper found that junior doctors bear high recovery costs and are the least able to defer requests from seniors.^19^ The present findings suggest a potential mechanism. The repair strategy that experimental work most clearly supports depends on standing relative to the interrupter, and that standing came from seniority or shared team membership. Regina, a consultant, lacked it with colleagues from outside her team. SIT accounts for this team boundary, which mirrors the boundaries between specialties that trainees have described elsewhere.^24^ Work on hierarchy and speaking up describes the seniority pattern.^36,37^ Reconstruction was also relational. Patients and parents helped doctors find their place, a form of partnership that eased resumption and depended on trust. It also carries a risk, because a patient or parent may recall the consultation inaccurately and a doctor who has lost the thread may not notice. Accumulated frustration also had to be managed before decisions, which suggests that models treating interruptions as discrete events may understate their cost.

### Organisational recognition may change the cost of interruption

The advice and informal supervision delivered through interruptions is what Star and Strauss call invisible work, essential to how a system functions but unrecorded within it and therefore unresourced.^41^ Its invisibility may also have cognitive consequences. Forewarning helps people prepare for an interruption and resume efficiently.^39^ Job planning may offer a similar forewarning at organisational level. Participants whose supervision was planned described its interruptions as expected work. Royal College of Radiologists guidance already recommends 0.25 sessions per week per trainee for formal clinical and educational supervision.^42^ The gap our participants described lay in unscheduled work, including ad hoc advice, vetting and telephone queries, and in clinic templates that were not reduced when a doctor was supervising.

### Implications

For practice, our findings favour coordination that is explicit and shared. Departments could agree thresholds for interrupting together with a named route for non-urgent requests, use designated and time-limited roles for fielding queries, reduce clinic templates for supervising doctors and count advice and vetting in job plans. These measures are intended to redirect non-urgent requests while keeping routes open for junior staff to escalate concerns. For research, interventions should be designed and evaluated at team and organisational level. Evaluations should ask who is able to use an intervention, as well as whether interruptions become less frequent. Rotating staff, cross-cover and short encounters are common across NHS secondary care, so these findings may apply beyond outpatient settings.

### Strengths and limitations

The study was designed around theory, and pairing a cognitive theory with a social one helped explain why strategies are felt to succeed or fail. Purposive sampling across experience and patient complexity, a multidisciplinary analytic team, double coding and participant validation strengthened credibility. The study also has limitations. All data were self-reported, so descriptions of strategies that worked reflect participants’ views and not measured effects. Participants often drew on work beyond the outpatient clinic, so strategies, such as the EPIC role, may need adapting before use in clinics. Few trainees took part, and most quotations came from consultants, so the range of trainee experiences may be underrepresented. We sampled only doctors, so the views of those who interrupt, including nurses, administrative staff and junior colleagues, were inferred from the accounts of those interrupted. Research that includes the whole team, and studies that test team and organisational strategies, are needed.

## Conclusion

Interruptions in outpatient care cannot be eliminated, and many should not be. Managing them well depends on how the cognitive cost of interruption is shared across teams, and on whether organisations recognise and legitimise that sharing. Interventions that target individual behaviour alone are likely to reach the ceiling our participants described. Shared norms, explicit roles and visible interruption work offer a sustainable way to regulate interruptions while keeping the ones that matter.

## Supporting information

Supplementary Materials referenced in text

## Data availability statement

No data are available. The ethics approval for this study does not permit the transcripts or other participant data to be shared.

## Declarations

### Contributors

SB conceived and designed the study, obtained ethical approval and funding, led data collection, contributed to the analysis and drafted the manuscript. CS contributed to the conceptualisation of the study, co-facilitated data collection, led the coding and analysis of transcripts, and contributed to drafting the manuscript. AR contributed to the conceptualisation of the study, independently double coded a subset of transcripts, provided supervision and critically revised the manuscript. AG contributed to the conceptualisation of the study, provided supervision and critically revised the manuscript. All authors approved the final version of the manuscript and agree to be accountable for all aspects of the work. SB is the guarantor and accepts full responsibility for the work and the conduct of the study, had access to the data, and controlled the decision to publish.

### Funding

This work was funded by the MPS Foundation, the charitable research arm of the Medical Protection Society. The MPS Foundation distributed the survey invitation through which participants were recruited. The research was conducted independently, and the funder had no role in study design, data collection, analysis or interpretation, the writing of the report, or the decision to submit the article for publication.

### Competing interests

None declared. No author is an employee of, holds a financial interest in, or is a member of the MPS Foundation or the Medical Protection Society.

### Ethics approval

This study involved human participants and was approved by the UCL Research Ethics Committee on 20 May 2025 (ID 1132). All participants gave informed consent before taking part.

### Patient consent for publication

Not applicable.

### Use of artificial intelligence

Claude (Anthropic), a generative AI tool, was used to help shorten and restructure the manuscript, draft revised text including the abstract and key messages, format it to journal requirements and check the reference list. The authors reviewed and edited all output, verified all content and references, and take full responsibility for the manuscript.

## Acknowledgements

We thank the doctors who gave their time to take part in this study.

## Notes

### Competing Interest Statement

The authors have declared no competing interest.

### Author Declarations

The UCL Research Ethics Committee gave ethical approval for this work (ID 1132, approved 20 May 2025).

