## Supplementary Materials referenced in text for "How should doctors manage workplace interruptions? A qualitative study in UK hospital outpatient departments"

**Contents**

- Online supplemental file 1. Sampling and participant validation
- Online supplemental file 2. Focus group and interview topic guide
- Online supplemental file 3. Full versions of quotations given in part in the Results

### Online supplemental file 1. Sampling and participant validation

#### Screener survey and stratification

The screener formed part of a national online survey of UK secondary care doctors (reference blinded for review). The survey invitation was emailed to the network of a UK medical defence organisation on 14 July 2025, with reminders on 28 July and 11 August, and the survey closed on 18 August 2025. Respondents could indicate interest in taking part in a focus group. The screener collected consent, role, specialty, years since qualification, self-reported interruption frequency (five-point scale from 1, low, to 5, high) and self-reported patient complexity.

Patient complexity was measured with a single item.

On a scale from 1 (low complexity) to 5 (high complexity), how would you rate the typical complexity of the patients you manage during consultations?

Response options 1 = Low, 2, 3, 4, 5 = High

Ratings were split at the median to give high and low complexity groups. In the full survey sample the median rating was 4 (n=415). Eligible respondents were then stratified into six groups by crossing experience level (trainee, 0–19 years post-qualification or 20+ years) with patient complexity (high or low). They were invited to sessions for their group so that focus groups were homogeneous for experience level.

#### Participant validation

A summary of draft findings, including the management themes reported in this paper, was shared with all participants. Eleven written responses and two non-verbal affirmations (‘thumbs-up’ reactions) were received. Ten written responses agreed that the findings resonated with their experiences. The remaining respondent also agreed, and added that interruptions have a cumulative effect, building up across a clinical session. This exercise supported the credibility of the findings and refined the final outputs.

#### Reference for online supplemental file 1

[Reference blinded for review]. National cross-sectional survey of UK secondary care doctors on interruptions. *medRxiv* [Preprint] 2026.

### Online supplemental file 2. Focus group and interview topic guide

*This semi-structured guide was used for online focus groups (grouped by similar levels of experience to mitigate power dynamics) and interviews. It is divided into sections exploring the nature of interruptions, their cognitive and social impacts, and strategies used for their management.*

**Opening and consent**

Hello, I’m [name] and I’ll be leading the interview/focus group today. Thank you for taking part. Before we begin I need to cover a few important ethical points.

Participation is voluntary and this session will take about 55 minutes. There aren’t any right or wrong answers and we’re just looking to get your opinions and experiences.

You’ve already given electronic consent via the screener; that consent covers your survey responses and the audio-recording for this focus group. If you would like to withdraw for any reason you can do so at any time during the session. Please note, however, that as transcripts will be pseudonymised, it will not be possible to remove contributions made to that point in the analysis.

Recordings and transcripts will be stored securely on [institution] systems, and only the research team will have access. We will also be taking notes during the session to remind us of important information.

If that’s all clear, I’ll start the recording. Do you have any questions or objections to being recorded?

**A. Introduction (5 minutes)**

- ‘As you know, we are interested in how unexpected interruptions, such as a colleague asking a quick question or a phone call in the middle of a consultation, affect your work. We also want to understand how these events might change the way you feel mentally, how they might influence interactions among team members, and how you manage an interruption once it’s occurred.’

**B. Experiences with interruptions (15 minutes)**

**C. Cognitive impacts and task demands (15 minutes)**

**D. Social dynamics and interpersonal aspects (15 minutes)**

**E. Management and coping strategies (10 minutes)**

‘Finally, I’d like to discuss the ways you’ve learned to cope with interruptions once they’ve happened.’

- ‘What strategies have you or your team adopted to manage interruptions effectively?’
  - *Prompt* ‘In what situations do your strategies work well, and when do they not?’
  - *Prompt* ‘How do you know which tasks to prioritise? Can you give an example?’
- ‘Have you used any structured approaches (eg, handover or communication protocols) that you believe could work better?’
- ‘What barriers prevent the effective management of interruptions in your setting? What factors or resources would facilitate better management?’

**F. Future directions and close (5 minutes)**

- ‘Based on your experiences, what improvements or interventions would you suggest to reduce the negative impact of interruptions?’

### Online supplemental file 3. Full versions of quotations given in part in the Results

Full versions of quotations that appear in part in the Results of the main manuscript. G, sampling group (see table 1 in the main manuscript). Ellipses (…) indicate omitted text.

| **Theme or subtheme** | **Quotation** |
| --- | --- |
| 1 Pre-emptive protection of attention | ‘I would literally go and secrete myself in a corner somewhere out of the way so it’s somewhere less findable, so I’m basically less distractible or less interruptible because I’m physically not there.’ (Dean, emergency medicine consultant, G4) |
|  | ‘I’ve even tried a hotel do not disturb thing on the door handle, but that doesn’t seem to deter people either.’ (Graham, ophthalmology consultant, G5) |
| 3.1 Anchoring before engaging | ‘If I’m midway through figuring something out and someone comes in, if I’ve got enough presence of mind, I’ll at least say to them please hang on while I finish typing the sentence. And that makes all the difference, because it means I can at least park my thoughts before they start speaking to me.’ (Jeanne, anaesthetics consultant, G3) |
|  | ‘Just let me finish off this task so that I can then concentrate fully because as someone’s already said if you’re doing something you’re not listening to it properly. Therefore, the advice you might give. And for a consultant it’s usually advice that you’re giving. That’s why you’re being interrupted, you’re being asked to do something or think about something and give them advice. I wouldn’t be able to do that properly unless I’ve actually fully concentrated on it, and therefore I can only do one thing at a time.’ (Victor, emergency medicine consultant, G3) |
| 3.2 Reconstructing the clinical narrative | ‘What I often do is try to recap what I was doing before … I say right, sorry for the interruption. So just to recap, you were saying that this started on Saturday and try to summarise back to them … just to give my brain time to like get back into what we’re doing.’ (Katrina, emergency medicine senior registrar, G2) |
|  | ‘I think, I suppose if it’s near the beginning, you’ve time to reflect. Yeah, maybe it’s more damaging at the end because you’re you’re nearer terminating the consultation, so there isn’t enough time to catch up … if it’s very near the beginning, you kind of reset and you come back and you can go. Where are we?’ (Ben, cognitive disorders specialist, G5) |
|  | ‘It’s difficult to reset yourself sometimes. Sometimes you just try and maybe a glass of water or coffee or something or go stretch your legs for a few minutes and then try and get back into it … I think being aware of it is important. So if I’m sort of aware that I’ve had, you know, another disruption, I’m quite frustrated. I’m aware that I’m frustrated and therefore try and take a few minutes to calm down before I go and validate the scan or something or make a definitive decision.’ (Ryan, radiology registrar, G2) |
| 4 Organisational recognition | ‘We didn’t have any protected time to actually go through education and clinic. They do it in a way that is chock a block. So you have enough time to see your own patient, but they expect you to do training on top of that.’ (Macy, consultant, G3)* |

*Specialty not stated in transcript.
